# Early Multidrug Outpatient Treatment of SARS-CoV-2 Infection (COVID-19) and Reduced Mortality Among Nursing Home Residents

**DOI:** 10.1101/2021.01.28.21250706

**Authors:** Paul E Alexander, Robin Armstrong, George Fareed, Kulvinder K. Gill, John Lotus, Ramin Oskoui, Chad Prodromos, Harvey A. Risch, Howard C Tenenbaum, Craig M. Wax, Peter A. McCullough

## Abstract

The outbreak of COVID-19 from severe acute respiratory syndrome coronavirus 2 (SARS-CoV-2) has spread all over the world with tremendous morbidity and mortality in the elderly. In-hospital treatment addresses the multifaceted nature of the illness including viral replication, cytokine storm, and endothelial injury with thrombosis. We identified nine reports of early treatment outcomes in COVID-19 nursing home patients. Multi-drug therapy including hydroxychloroquine with one or more anti-infectives, corticosteroids, and antithrombotic agents can be extended to seniors in the nursing home setting without hospitalization. Data from nine studies found multidrug regimens relying on the use of hydroxychloroquine as well as other agents including doxycycline were associated with a statistically significant and >60% reductions in mortality. Going forward, we theorize and based on the evidence, that early empiric treatment for the elderly with COVID-19 in the nursing home setting (or similar congregated settings with elderly residents/patients) has a genuine probability of success and acceptable safety. This group remains our highest at-risk group and warrants acute treatment focus that will prevent the development and/or worsening of problems associated with COVID-19, most particularly isolation, hospitalization, and death. In fact, with the rapidity and severity of SARS-CoV-2 outbreaks in nursing homes, in-center treatment of patients with acute COVID-19 is possibly the most rational and importantly *feasible* strategy to reduce the risks of hospitalization and death. If the approach remains ‘wait-and-see’ and elderly high-risk patients in such congregated nursing room type settings are allowed to worsen with no early treatment, they may be too sick and fragile to benefit from in-hospital therapeutics and are at risk for pulmonary failure, life-ending micro-thrombi of the lungs, kidneys etc. We put forth the notion that the most important factor in this regard, is making available early therapeutic intervention as described here. These drugs include and under supervision by skilled doctors, combination/sequenced ivermectin, hydroxychloroquine, colchicine, azithromycin, doxycycline, bromhexine hydrochloride, and favipiravir (outside the US), along with inhaled steroids such as budesonide and oral steroids including dexamethasone and prednisone, and anti-thrombotic anti-clotting drugs such as heparin). As the clinical trials data on treatments for COVID-19 mature, this early treatment therapeutic option deserves serious, urgent, and sober consideration by the medical establishment and respective decision-makers.

## Background

The pandemic of SARS-CoV-2 virus and the resulting clinical disease known as COVID-19, has spread across the world relentlessly and it appears now that it is becoming endemic. This pathogen demonstrates high infectivity while the pathogenicity at present continues generally to be low. Yet it still causes devastating consequences to a proportion of high-risk persons, principally the elderly among us, and in particular those with underlying medical conditions oft referred to as ‘co-morbidities’. Hence, we recognize and appreciate that COVID-19 is a devastating illness for elderly persons with underlying medical conditions who are infected. Case fatality (mortality) rates are elevated and have been based thus far on laboratory confirmed cases and have not yet included an accurate reflection of infection, for example mild or asymptomatic cases that have recovered. Ideally, a more accurate and reliable infection fatality rate should be reported as comprehensive data become available which would provide a more precise reflection of lethality.

In the previous year, we have found that SARS-CoV-2 may lead to a spectrum of COVID-19 syndromes including but not necessarily limited to asymptomatic exposure, symptoms of the common cold, influenza-like symptoms, as well as fulminant multiorgan system failure, the latter being responsible in large part for most fatalities that have occurred. The most important variable for risk stratification for both hospitalization and death is advanced age, which explains why nursing home patients have accounted for a large fraction of deaths in most developed countries.

A policy brief published by the American Geriatrics Society (AGS) has outlined clearly the impact of COVID-19 pathophysiology and severity of illness in the nursing home, highlighting the grave challenge in treating and preventing death in this patient population. The AGS reported that over 15,000 nursing homes and long-term care facilities provide care for the oldest Americans who are at greatest risk for COVID-19 and its complications (pulmonary failure and death) especially as they have multiple comorbidities.^1,2^ The AGS suggests consideration of ‘hospital-at-home’ care models as a practical path forward.

Unfortunately, in nursing home residents there have been no large, well-funded, high-quality randomized trials of single or multidrug regimens for COVID-19. Given the enormous public health importance of the issue, we sought to assemble the available information concerning treatments that have been attempted in senior homes (nursing homes, old-aged homes, care homes, long-term care homes, assisted-living facilities etc.) and their associated outcomes. In most circumstances, the comparator or approach was watchful waiting and then hospitalization for severe symptoms for those not afforded early multidrug treatment.

## Methods

For pragmatic reasons we searched PubMed/MEDLINE database for literature relevant to the question addressed here. Our literature search spanned a period up to January 2021 using relevant search terms including COVID-19, SARS-CoV-2, nursing homes, long-term care, nursing home residents, high-risk, elderly residents, mortality, death, treatment, and early treatment, to identify reports of attempts to treat COVID-19 in nursing homes with the primary outcome of study being hospitalization and/or mortality. We sought to access as complete a body of evidence as appropriate to inform this manuscript. Thus, study reference lists were also hand searched for any potentially eligible reports. Evidence was also considered, where available, from additional sources such as online preprint publications not yet having completed the peer-review process (e.g. clinical medicine preprint repository, medRxiv.org etc.). This said, we point out that such as yet unreviewed papers were subjected to rigorous review by the authors before being considered for inclusion in this paper.

Relevant reports were examined in duplicate and independently for full agreement on final eligibility to inform the review. We were prepared to discuss in consensus debate in instances of potential eligibility disagreement and use of adjudication if necessary. From the reports, we extracted data on mortality among those treated with one or more drugs against COVID-19 and also for those in comparator groups. This step was also conducted in duplicate and checked for accuracy. MedCalc (https://www.softpedia.com/get/Science-CAD/MedCalc.shtml) was used to calculate odds ratios/relative risks and their 95% confidence intervals and p-values for mortality. For the purposes of this review, we defined ‘nursing home’ as any nursing home, long-term care facility, care home, assisted-living facility.

In sum, our approach towards inclusion of references in this manuscript was as stringent as possible and included the same systematic and methodological rigor reflective of approaches used for systematic reviews. We sought to alert the clinical and medical research community as well as policy makers, with the accumulated benefits of early treatment in the elderly population that remains at greatest risk of hospitalization and death from COVID-19. Our cardinal aim therefore given the accumulating evidence, is a summons to the medical research community for urgent high-quality, trustworthy, and robust comparative effectiveness research on early treatment in high-risk and symptomatic SARS-CoV-2 positive patients/residents (especially in the nursing home environment). Having said this, we also implore the medical community to consider that whilst in the midst of this terrible pandemic, we should not await, nor is there a need to await the completion of randomized controlled trials. Indeed, based on the data we present here we would suggest that we are bound ethically to proceed now with treatment that has been shown to be effective by way of other highly acceptable clinical study methods.

## Results

As a result of our search methods, we found 336 initial reports that were potentially eligible for our review. However, after applying the scrutiny and stringency of reference analysis described above we judged that nine (9) studies met our criteria for inclusion (a result not dissimilar to most systematic reviews that have been published we might add).^3-11^ These shed light on the impact of early outpatient sequential multidrug treatment (SMDT^12^) in nursing home residents (Table 1). We refer here to therapeutics that are provided under clinician/health expert/provider supervision and in a sequenced manner with carefully considered dosing and timing and direct the reader to *Reviews in Cardiovascular Medicine* for complete details on the suggested early outpatient ambulatory SMDT dosing and treatment algorithm with combinations of well-known, safe, cheap, and effective drugs. These drugs include ivermectin, hydroxychloroquine, colchicine, azithromycin, doxycycline, and favipiravir (outside the US), along with inhaled steroids such as budesonide and oral steroids including dexamethasone and prednisone, and anti-thrombotic anti-clotting drugs such as heparin^12^). We also present the relative reduction in mortality risk (Figure 1) where data from certain selected studies described below allowed such computations.

**Table 1:** Reports of early prehospital SMDT in nursing home residents

| First author's surname | Treatment | Outcome |
| --- | --- | --- |
| Bernabeu-Wittel<sup>3</sup> | HCQ/HCQ + lopinavir/ritonavir/HCQ + AZM | Survival in active treatment group independently associated with use of any antiviral treatment; survival in the antiviral treatment group was independently associated with receiving any of the antiviral treatments (OR = 28 [5-160]). |
| Heras<sup>4</sup> | HCQ/HCQ + AZM | HCQ + AZ correlated significantly with decreased mortality, HCQ alone also correlated but not significantly; 83 received pharmacological treatment, total survival was 76%, 7 (8%) patients were hospitalized, of which 4 died. |
| De Spiegeleer<sup>5</sup> | Effect of medication for preexisting conditions: Angiotensin-converting enzyme inhibitors (ACEi), angiotensin II receptor blockers (ARBs), or HMB-CoA reductase inhibitors (statins) | Statins significantly correlated with decreased mortality, ACEi and ARBs also correlated but not significantly; a statistically significant relationship between statin intake and the absence of symptoms during COVID-19 (OR 2.91; 95% CI 1.27 to 6.71). |
| Brouns<sup>6</sup> | Effect of medication for preexisting conditions: Oral antithrombotic therapy (vitamin K antagonists, direct oral anticoagulants, anti-platelet therapy) | Association of antithrombotics with diminished mortality but not significant and further reduced when controlled for age, sex, hypertension, and comorbidity; univariate analysis (OR 0.89, 95% CI 0.41 to 1.95, p=0.776). |
| Ahmad<sup>7</sup> | HCQ + DOXY | Smaller percentage of deaths and hospitalizations compared to a demographically similar nursing home where residents were not treated; of 54, 9 did not complete treatment, and those completing the course of treatment were afebrile by at least 5 days after completion, and either had no more shortness of breath or had returned to previous oxygenation levels (for patients who had been on ventilation before contracting COVID-19). |
| Ly<sup>8</sup> | HCQ + AZM | Treated residents had significantly lower mortality. The overall mortality was 20.8%, and hospitalization including ICU admission was 20.8%. Mortality was lower in patients treated with HQC + AZM for at least 3 days |
|  |  | (15.5%) than in those not so treated (26.4%), OR 0.37, p = 0.02. |
| Alam<sup>9</sup> | DOXY + standard of care | Researchers concluded that early treatment with DOXY for high-risk patients with moderate to severe COVID-19 infections in non-hospital settings, such as LTCFs, is associated with early clinical recovery, decreased hospitalization, and decreased mortality; 85% of patients (n=76) revealed clinical recovery, 3% hospitalized |
| Cangiano<sup>10</sup> | HCQ/heparin/corticosteroid/anti-platelet | Mortality was significantly lower for patients who had previously been on chronic vitamin D supplementation, and was significantly reduced by COVID-19 treatment with HCQ (p=0.03, 28 survived vs 5 who died). |
| Leriger<sup>11</sup> | HCQ/non-HCQ | After 2 weeks, 76 (72%) of residents receiving HCQ showed no symptoms, while 9% had symptoms, 2% continued in hospitalized, and 17% had died; control group had 58% (66) who were asymptomatic, 13% who had mild symptoms, 1% with moderate symptoms, 3% hospitalized, and 25% had died; the benefits of HCQ treatment in positive nursing home residents seem to outweigh the harms in terms of symptom severity and mortality, and particularly so in residents over 80 years of age and when HCQ was administered before symptoms began (started early). |

**Figure 1:**
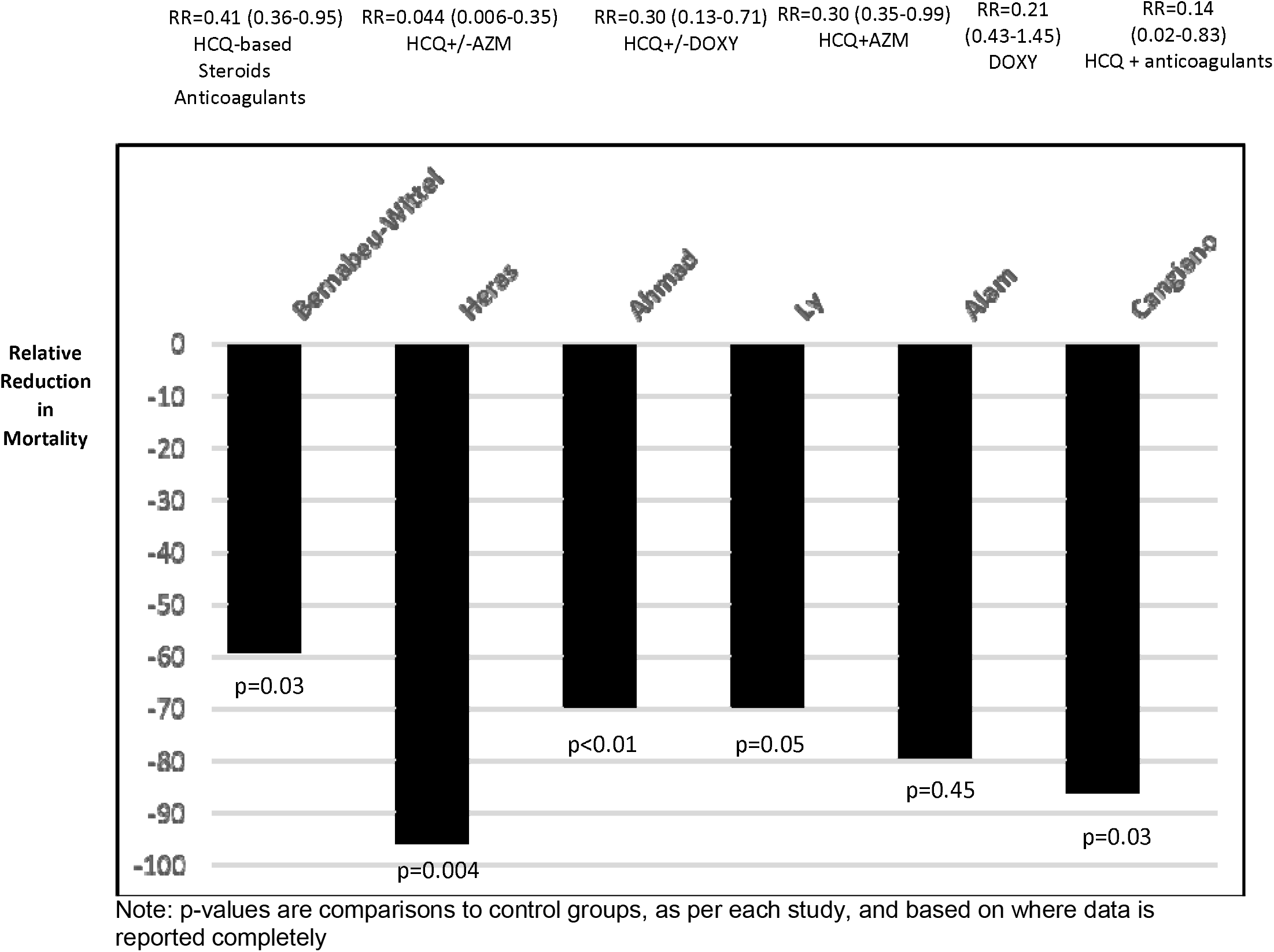
Relative reduction in mortality risk in nursing home COVID patients using early prehospital SMDT

Bernabeu-Wittel et al. reported from four nursing homes in Spain^3^ that 272 of 457 (59.5%) residents contracted SARS-CoV-2 of whom 189 (69.5%) were given ‘active standard care’ and the rest were given advanced palliative care. For patients assigned to active standard care (i.e. *medicalization*, see Table 1), survival was measured as both dichotomous and time-dependent outcomes. The findings demonstrated that 139 (73.5%) of these patients received antiviral treatment: 114 (60%) received hydroxychloroquine (HCQ), 18 (10%) HCQ + lopinavir/ritonavir, 7 (5%) HCQ + azithromycin (AZM). The investigators reported that survival in the ‘antiviral’ treatment group was associated independently with receiving any of the antiviral treatments (OR = 28 [5-160]). In addition, 119 (44%) received low molecular weight heparin, 62 (23%) and also received antimicrobials (e.g. “mild/moderate: oral amoxicillin/clavulanic acid, levofloxacin, or AZM; severe: parenteral ceftriaxone plus levofloxacin (or AZM; possibility of aspirative origin: oral amoxicillin/clavulanic acid, or parenteral ertapenem”), and 57 (21%) received systemic corticosteroids. Fewer patients who received active standard care (i.e. medicalized treatment) died (24 (13%)). The researchers also reported that the ‘medicalization’ program (as opposed to watchful observation) led to increased survival in this group, 82% before MP, 96% during MP (p = 0.004). Survival time in actively treated patients was associated independently with use of any antiviral treatment. Hospital referrals were also recorded, but for all patients, including those who did not receive active care; hospitalization significantly decreased with introduction of the medicalization program (Table 1).

Heras et al. reported on COVID-19 patients from a single nursing home in Spain.^4^ This was designed as a retrospective cohort case-controlled analysis. There were 100 confirmed cases and a medicalization approach to treatment had been carried out in 83 subjects. It was reported that most, 70%, received HCQ and AZM while others received variously HCQ alone and in some cases beta-lactam or quinolone antibiotics. Total survival was 76%, 7 patients were hospitalized, of which 4 died. No statistically significant improvement in survival was observed for patients who started treatment within the first 24 hours. Significantly greater risk of mortality was observed for lack of pharmacological treatment and type of treatment. Of survivors, 81.6% had received HCQ + AZM, whereas only 33.3% of those who died had received it. Of survivors, 7.9% had received HCQ alone, while 12.5% of those who died had received it. Of survivors, 10.5% had received ‘no treatment’, which also included beta-lactam or quinolone antibiotics, while 54.2% of those who died received it. No descriptions of the treatment for the 7 hospitalized patients were provided. In adjusted models, HCQ + AZM conveyed a 22.6-fold lower mortality risk, p=.004 (Figure 1) compared to use beta-lactam or quinolone antibiotics only.

De Spiegeleer and coworkers published outcomes from two Belgian nursing homes in 2020^5^ to analyze the association between angiotensin converting enzyme inhibitor (ACEi)/angiotensin receptor antagonist (ARB) and/or statin use with the clinical outcome of COVID-19 (n=154 COVID-19-positive subjects). Twenty percent were taking ACEi/ARBs and 20% were taking a statin, while 5% were taking both. Researchers reported a statistically significant relationship between statin intake and the absence of symptoms during COVID-19 (OR 2.91, 95% CI 1.27 to 6.71), retaining significance when adjusted for covariates. Forty-seven percent of 154 SARS-CoV-2 positive patients remained asymptomatic and 24% had severe disease. The conclusion was that statin use in such older and vulnerable nursing home residents is associated potentially with a significant positive effect (better outcome) on COVID-19 clinical symptoms and course. The authors also reported that “The fact that statin intake is more strongly associated with asymptomatic status than serious COVID-19 suggests that the potential therapeutic effects of statins are more outspoken (*sic*) in the initial stages of COVID-19.”^5^

Broun et al. reported a retrospective case series involving 14 nursing homes in the Netherlands (n=101 residents),^6^ concerning oral anticoagulants and mortality in nursing home residents with COVID-19. The overall mortality was 47.5% (48 deaths). Researchers found that anticoagulation was related to non-significant reduction in mortality based on univariate analysis (OR 0.89, 95% CI 0.41 to 1.95, p=0.78) with adjustment for gender, age, hypertension, and comorbidities.

Ahmad et al. published a report of 52 confirmed SARS-CoV-2 positive COVID-19 patients^7^ from three nursing homes in New York. Upon diagnosis, residents were treated with doxycycline (DOXY) and HCQ. DOXY reportedly acts as a broad-spectrum inhibitor of MMPs (potentially having a direct action on matrix-metalloproteinase-mediated inflammation/tissue damage) and also acts as an antioxidant.^13-16^ Additionally, researchers indicate that tetracyclines such as DOXY can impact viral infections via their direct antibacterial properties as well as via direct antiviral activity, thus capable of slowing/inhibiting COVID-19 disease progression^17^ as has also been shown in other human inflammatory diseases.^17,18^ Patients were followed for 11 days with treatment course of 7 days.^7^ Nine patients did not complete the 7-day therapy, 6 due to hospitalization, 2 due to death in the nursing home, and 1 due to adverse reaction (seizure). At the time of publication, of these 9, 3 had died. The remaining patients completing the course of treatment, were afebrile by at least 5 days after completion, and either had no more shortness of breath or had returned to previous oxygenation levels (for patients who had been on oxygen supplementation before contracting COVID-19). Researchers concluded that treatment with DOXY-HCQ in high-risk COVID-19 patients was associated with a benefit in clinical recovery, decreased transfer to hospital and decreased mortality.^7^ For comparison, the authors also reported on a nursing home with similar demographics in Washington whose residence did not receive pharmacological treatment and had 57% hospitalization and 22% mortality, as compared to 11% and 6% in the study group, respectively.

Ly et al.^8^ reported on the results of SARS-CoV-2 PCR-based screening campaigns conducted on dependent elderly residents (compared with staff members) in long-term care facilities (LTCFs) in Marseille, France, with follow-up of positive cases. This was a retrospective study of positive residents in 24 nursing homes whereby 226 of 1,691 residents (13.4%) tested positive. In total, 116 (51.4%) patients received courses of oral HCQ and AZM for ≥ 3 days and these patients otherwise differed little in comparison to patients who did not receive the treatment. The overall mortality was 20.8%. Mortality was reportedly lower in patients treated with HCQ + AZM for at least 3 days (15.5%) than in those not so treated (26.4%), adjusted OR = 0.37, p = 0.02. Researchers concluded that the elevated proportion of asymptomatic COVID-19 patients and independent factors for mortality “suggest that early diagnosis and treatment of COVID-19 patients in long-term care facilities (LTCFs) may be effective in saving lives”.^8^

Alam and coworkers ^9^ conducted a retrospective case-series study in New York to assess and document clinical outcomes of high-risk COVID-19 LTCF patients (n=89 persons diagnosed March to May 2020). These patients had early intervention with doxycycline (DOXY) after presenting with moderate to severe symptoms. All 89 had developed sudden onset of fever, cough, shortness of breath (SOB), and hypoxia. Treatment with DOXY began within 12 hours of symptom onset, 100 mg bid PO or intravenous (IV) for seven days and regular standard of care (11 additionally received broad-spectrum antibiotics). Researchers reported that 85% of patients (n=76) had clinical recovery, “defined as resolution of fever (average 3.7 days, Coeff = −0.96, p = 0.0001), resolution of SOB (average 4.2 days), and improvement of POX: average 84% before treatment and average 95% after treatment (84.7 ± 7% vs. 95 ± 2.6%, p = 0.0001). Higher pre- and post-treatment POX is associated with lower mortality (oxygen saturation (Spo2) vs. Death, Coeff = −0.01, p = 0.023; post-Spo2 vs. Death, Coeff = - 0.05, p = 0.0002)”.^9^ Ten patients died (11%) within 10 days of symptom onset and 3% were transferred to hospital. Researchers concluded that “early treatment with DOXY for high-risk patients with moderate to severe COVID-19 infections in non-hospital settings, such as LTCFs, is associated with early clinical recovery, decreased hospitalization, and decreased mortality”. The data from Alam et al.^9^ was compared to similar age groups in New York (Yang et al. 2020^19^) during the same period of the pandemic in order to calculate the relative risk reduction with DOXY.

Cangiano et al.^10^ studied mortality of nursing-home COVID-19 patients in Milan, Italy using an observational study design. These patients, average age 90 years, were treated in the facility with a standard-of-care multidrug therapy including HCQ, corticosteroids, and antithrombotics. Of the 98 patients followed, 56 survived and 42 died over the two months of the study. The authors found that mortality was significantly lower for patients who had previously been on chronic vitamin D supplementation, and was significantly reduced by COVID-19 treatment with HCQ (p=.03).

Finally, Leriger et al. (2020)^11^ followed 233 residents in 11 skilled nursing homes in Indiana who had tested positive for COVID-19 (113 residents who acted as controls were selected from the same nursing homes and also followed for 14 days (all testing positive). Hydroxychloroquine (HCQ) was administered to 105 nursing home residents (45%). After 2 weeks, 76 (72%) of residents receiving HCQ showed no symptoms, while 9% had symptoms, 2% continued in hospitalized, and 17% had died. The control group had 58% (66) who were asymptomatic, 13% who had mild symptoms, 1% with moderate symptoms, 3% hospitalized, and 25% had died. The 2 residents who were hospitalized in the HCQ group recovered (initially hospitalized for chest pain/tachycardia) and asymptomatic after 2 weeks. Researchers concluded that the benefits of HCQ treatment in nursing home residents who are COVID-19 positive, seem to outweigh the harms in terms of symptom severity and mortality, and particularly so in residents over 80 years of age and when HCQ was administered before symptoms began (started early).

## Discussion

We found nine reports^3-11^ supporting the concept of early multidrug pharmacological intervention can be used to improve clinical outcomes (the most important being reduction in death) in elderly nursing home residents suffering from COVID-19. We were focused on a high-risk group that typically worsens before treatment is administered within an in-patient hospital setting. In essence, the reports from nursing homes showed that early multidrug interventions including most commonly two or more intracellular anti-infectives, and before hospitalization, were associated with overall reductions in mortality of > 60%. These findings, although derived from independent investigations, were externally consistent and in most instances, clinical improvements as well as reduced rates of mortality were shown to be statistically significant and clinically significant. While the reports focused on HCQ and anti-infectives, as indicted in a prior early outpatient SMDT study,^12^ early treatment can potentially include other antivirals/anti-infectives, corticosteroids, and anti-thrombotic/platelet drugs based on clinically valid decision-making. Our data imply that early medical therapy in addition to several other interventions in nursing homes when combined could reduce hospitalization rates dramatically and improve survival.

There can be no doubt that the COVID-19 crisis has markedly increased annualized death rates in nursing home. For example, an Italian observational study reported a two-month mortality of 40%, compared to 6.4% in the prior year (COVID-19 positive residents (43% increase) and negative residents (24% increase).^10^ Greater mortality was associated with being male, older, no previous vitamin D supplementation and lower “activities of daily living (ADL)” scores,^10^ leaving researchers to conclude that there is a greater elderly mortality due to COVID-19.

Similarly, Panagiotou et al. (2020)^20^ sought to identify risk factors for 30-day all-cause mortality among US nursing home residents with COVID-19. The study was conducted in 351 US nursing homes involving 5256 nursing home residents with COVID-19–related symptoms who had severe ARDS precipitated by SARS-CoV-2 infection that was confirmed by RT-PCR testing (March to September). The median age was 79 years (IQR, 69-88 years) and researchers reported that increased age, being male, and weakened cognitive and physical function among these US nursing home residents with COVID-19 were independently associated with all-cause 30-day mortality.

Nursing home staff play a critical role in spreading and potentially containing the SARS-CoV-2 outbreak. A report from France^21^ involving 17 nursing homes with 794 staff members confined to the facility along with 1,250 residents were compared with a national sample of 9,513 facilities with 385,290 staff members and 695,060 residents. Lower rates of COVID-19 were found among facilities that had staff members live in the facility as compared to those who lived at home and traveled to the nursing home for work each day. These findings suggest SARS-CoV-2 is spread from families (living in the same home) of nursing home workers to the nursing home residents who were in lockdown. Similarly, Davidson and Szanton (2020)^22^ called for better management of nursing home residents in the era of COVID-19 with reductions in overcrowding and congregate settings. Additionally, researchers^23^ reported that outbreaks of COVID-19 are associated with the size of the nursing home type facility whereby institutions with > 150 beds will experience a much greater likelihood of transmission of the virus compared to smaller facilities.

Surveillance for SARS-CoV-2 is an essential strategy for evaluating SARS-CoV-2 transmission within nursing homes. Along these lines of greater risk in nursing home settings, Graham et al. (2020)^24^ conducted an outbreak investigation of 394 residents and 70 staff in 4 nursing homes impacted by COVID-19 outbreaks in central London. The investigation found that 26% of residents died over the two-month period and all-cause mortality increased by 203% compared to the same period in prior years. Testing identified 40% of residents as positive for SARS-CoV-2 (43% of them asymptomatic and 18% had only atypical symptoms with 4% of asymptomatic staff also testing positive). Similarly, researchers (2020)^25^ reported very elevated death (33% of residents (34/101)) from COVID-19 in a long-term care facility in King County, Washington. After detecting one infected resident late February, by mid March, there were 167 confirmed cases of COVID-19 involving 101 residents, 50 health care personnel, and 16 visitors. Researchers reported that hospitalization rates for residents, visitors, and staff were 54.5%, 50%, and 6%, respectively. Similarly, by this time in March, 30 long-term care facilities were reporting at least one confirmed case of COVID-19.

Likewise, a recent report on Singapore’s handling of COVID-19^26^ showed that residents in nursing homes and long-term care facilities are at greater risk of transmission and severe illness and deaths. While Singapore’s nursing homes and long-term care facilities accounted for <0.001% of total COVID-19 cases in April 2020, they accounted for 14% of all deaths from COVID-19. Arons et al. (2020)^27^ sought to assess transmission risk after one case of COVID-19 emerged in a nursing facility in Washington. They found elevated transmission and death from that *single* initial case whereas at 23 days from the initial positive case, 64% of the 89 residents tested positive, 11 were hospitalized (19%) and 15 died (26%). Approximately 99% of residents who were evaluated had at least one co-existing condition. A report on the COVID-19 situation in nursing homes in Hungary^28^ echoed very similar findings on the significantly elevated risk of transmission in such settings, estimating that 42% to 57% of all COVID-19 deaths occurred in nursing homes in European nations such as Italy, Spain, France, Ireland, and Belgium.

SARS-CoV-2 can spread quickly in nursing homes. A report on the COVID-19 in England and Wales^29^ discussed the rapidity at which the infection spreads in such settings, and it was reported that by the time the first resident developed symptoms, approximately 50% of the residents had already been infected. The UK Office for National Statistics (ONS) reported that in April, 72% of deaths in care homes were related in some way to COVID-19. These findings unlike the SARS-CoV-1 coronavirus epidemic in 2003,^30^ indicate that contagion-control methods alone are inadequate for COVID-19 and immediate treatment is needed to reduce mortality.

### Therapeutic Nihilism

Reports from multiple sites in the world indicate that therapeutic nihilism is practiced with seniors who do not have access to healthcare outside of their facilities. This is indeed very distressing if there are potential treatment options. It has been reported that in Quebec, Canada, as of December 2, 2020, 63% of deaths due to COVID-19 occurred in private and public nursing homes (4,493 of 7,097).^31,32^ As of January 9, 2021, it has been reported that in Ontario, Canada, 60% of deaths due to COVID-19 occurred in private and public nursing homes (approximately 3,000 of nearly 5,000).^33^ In The Chief Public Health Officer of Canada’s Report on the State of Public Health in Canada 2020 and looking specifically at COVID-19 in Canada, it was reported that 80% of all COVID-19 deaths in Canada were associated with nursing homes.^34^ Importantly and what we are arguing against, is that residents who are ill with COVID-19 are not provided any form of existing safe, simple, and inexpensive therapeutic drugs.^31^

The Centers for Disease Control and Prevention (CDC)^35^ recently highlighted the COVID-19 vulnerability among residents and staff members in LTCFs, and focused specifically on assisted living facilities (ALFs). LTCFs include ALFs and similar residential facilities, skilled nursing facilities (SNFs) as well as other nursing homes, and also include residential facilities for residents with varying degrees of intellectual and developmental disabilities. As of early November 2020, approximately 91,500 deaths were reported among residents and staff in LTCFs within the United States, representing 39% of total state COVID-19 deaths. By mid October 2020 and based on available ALFs data from 39 US states, among the 28,623 ALFs, 22% reported at least one COVID-19 case in their staff or residents. In addition, the cases that died comprised 21.2% of the residents and 0.3% of staff. This report made no mention of treatment provided to staff or residents. A print media report ^36^ stated that almost 50% of US COVID-19 deaths were linked to nursing homes. In addition, while the nursing home death toll was elevated significantly in New York, the deaths in New York’s nursing homes have been much higher than the state data reported.^37^ None of these reports indicate that nursing home patients have been offered any forms of prehospital therapy.

### Limitations of This Study

Our report has all the limitations of reviews that extract data from multiple heterogeneous studies. We did focus on nursing home residents who are usually weaker and more infirmed that persons living independently in the community, making our results not applicable to community-dwelling adults. The reporting of early treatment in COVID-19 patients can be considered nascent and thus the number of available studies is limited as is their sample sizes. We relied on data presented in study reports, which in non-randomized studies may be subject to variable degrees of adjustment for potentially confounding factors. We assumed that the various studies and their study subjects were sufficiently similar to draw conclusions across them, and that the care facilities examined in the study reports are typical of such facilities in general and thus that the treatment benefits that we observed apply to nursing home and similar care facilities quite generally.

## Conclusions

In conclusion, nursing home residents are at the highest risk of death from SARS-CoV-2 infection and the development of COVID-19 illness and they appear to be the victims of spread from staff who live in the community. The available reports indicate there is a large > 60% mortality risk reduction associated with multidrug treatment regimens that utilize two or more intracellular anti-infectives (HCQ and either AZM or DOXY) combined with other agents including corticosteroids, anti-thrombotics (anti-platelets), and nutraceuticals of > 60%!^12^ There is also recent focus on the anti-parasitic ivermectin^38-40^ as part of early outpatient treatment, functioning as an antiviral/anti-infective. Ivermectin is less known in the West and is widely prescribed in Africa and Asia to treat parasitic diseases such as lymphatic filariasis or onchocerciasis (river blindness). Recent evidence including clinical reports, suggests it may reduce mortality during treatment of COVID-19, especially in patients with severe pulmonary involvement. Additionally, favourable evidence is accumulating regarding the potential anti-viral role of bromhexine hydrochloride (mucolytic cough suppressant that is a TMPRSS2 inhibitor that may function to block viral entry into the cell) in reducing the ICU transfer, intubation, and the mortality rate in patients with COVID-19.^41,42^

We recognize that large randomized, placebo-controlled, multidrug clinical trials are needed and will provide conclusive evidence in the future on the efficacy of early treatment of COVID-19. However, based on the reporting and clinical experience, the early therapeutics that we advocate for such as HCQ, ivermectin, and doxycycline etc.^12,17, 18, 38, 39^) may potentially function by working to inhibit viral replication while also dampening the host innate immune response thereby reducing oxidative stress, matrix-metalloproteinase-mediated inflammation/tissue damage, as well as the production of a wider array of inflammatory cytokines. In the meantime then and given the dearth of such randomized evidence that would provide clarification, and given the present emergency crisis, the observational non-randomized data we reviewed which we judged as acceptable to informing this report (and in line with expert arguments for observational study evidence use especially when RCT evidence is not available or is in many cases, of poor methodological quality),^43^ suggest a treatment mandate is present and should replace therapeutic nihilism for nursing home residents with acute COVID-19. In addition, observational research evidence is indeed evidence, and is an important component that also informs evidence-based decision-making. Indeed, the ‘21st Century Cures Act’^44^ was passed and specifically section 3022 addresses the use of observational research as part of medical research. Our goal is to save lives in an emergency. Furthermore, using a therapeutic outside of its authorized use is well acceptable if it is safe, used under expert prescribing and supervision, and showing success as a repurposed treatment in a crisis situation.

There are currently treatments which, applied from the first symptoms of COVID-19, have a very reasonable chance of mitigating the severity and duration of the symptoms, and of reducing the risk of hospitalization, death, or even sequelae. We have thus concluded that it is not possible to overstate the philosophy that since early in-center treatment with already available effective, safe, and relatively cheap medications (repurposed) in nursing homes is associated with a large reduction in mortality among nursing home residents, there can be no scientifically sound reasons, nor moral rationale for not utilizing these forms of treatment. In the urgent quest to save lives and particularly of the elderly nursing home infected resident, we urge clinical trials and advanced practice to immediately pivot to combination/sequential drug therapy for ambulatory COVID-19 illness. We urge that doctors be made aware of these treatment options and discuss them with their patients who are at risk of starting COVID-19, so that patients are fully informed of the options. The accumulating early treatment evidence is compelling and deserving of very serious and sober consideration as a therapeutic option given this emergency and the pressing morbidity and mortality. To do otherwise is to fail our patients, and to suggest placebo when there is data of favourable effects of early treatment is becoming more and more problematic to accept.^45^ We must never forget that patients have legal rights to be in receipt of effective treatments once they exist. We ask that you do not discount early treatment without your careful reflection, debate, and clinical discretion on offering early treatment at this emergency time period. Senator Ron Johnson of the US Senate held recent hearings to discuss COVID-19 and the early outpatient treatment option^46,47^ and we ask that his lead be followed up urgently given the urgency of the situation.

## Data Availability

All data can be provided by Dr. Peter McCullough;

## Acknowledgement

We thank Mr. Erik Sass, editor-in-chief of The Economic Standard for his oversight and input.

## References

1. American Geriatrics Society. American Geriatrics Society Policy Brief: COVID-19 and Nursing Homes. J Am Geriatr Soc. 2020 May;68(5):908–911. doi: 10.1111/jgs.16477. Epub 2020 Apr 29. PMID: 32267538; PMCID: PMC7262210.

2. Centers for Disease Control and Prevention (CDC). People who are at higher risk for severe illness. url: https://www.cdc.gov/coronavirus/2019-ncov/need-extra-precautions/people-at-higher-risk.html. (Accessed on December 25th 2020).

3. Bernabeu-Wittel M, Ternero-Vega JE, Nieto-Martín MD, Moreno-Gaviño L, Conde-Guzmán C, Delgado-Cuesta J, Rincón-Gómez M, Díaz-Jiménez P, Giménez-Miranda L, Lomas-Cabezas JM, Muñoz-García MM, Calzón-Fernández S, Ollero-Baturone M. Effectiveness of an On-Site Medicalization Program for Nursing Homes with COVID-19 Outbreaks. J Gerontol A Biol Sci Med Sci. 2020 Aug 1:glaa192. doi: 10.1093/gerona/glaa192. Epub ahead of print. PMID: 32738140; PMCID: PMC7454360.

4. Heras E, Garibaldi P, Boix M, Valero O, Castillo J, Curbelo Y, Gonzalez E, Mendoza O, Anglada M, Miralles JC, Llull P, Llovera R, Piqué JM. COVID-19 mortality risk factors in older people in a long-term care center. Eur Geriatr Med. 2020 Nov 27:1–7. doi: 10.1007/s41999-020-00432-w. Epub ahead of print. PMID: 33245505; PMCID: PMC7693854.

5. De Spiegeleer A, Bronselaer A, Teo JT, Byttebier G, De Tré G, Belmans L, Dobson R, Wynendaele E, Van De Wiele C, Vandaele F, Van Dijck D, Bean D, Fedson D, De Spiegeleer B. The Effects of ARBs, ACEis, and Statins on Clinical Outcomes of COVID-19 Infection Among Nursing Home Residents. J Am Med Dir Assoc. 2020 Jul;21(7):909-914.e2. doi: 10.1016/j.jamda.2020.06.018. Epub 2020 Jun 15. PMID: 32674818; PMCID: PMC7294267.

6. Brouns SH, Brüggemann R, Linkens AEMJH, Magdelijns FJ, Joosten H, Heijnen R, Ten Cate-Hoek AJ, Schols JMGA, Ten Cate H, Spaetgens B. Mortality and the Use of Antithrombotic Therapies Among Nursing Home Residents with COVID-19. J Am Geriatr Soc. 2020 Aug;68(8):1647–1652. doi: 10.1111/jgs.16664. Epub 2020 Jul 21. PMID: 32633418; PMCID: PMC7361386.

7. Ahmad et al. 2020. Pre-print. Doxycycline and Hydroxychloroquine as Treatment for High-Risk COVID-19 Patients: Experience from Case Series of 54 Patients in Long-Term Care Facilities. url: https://www.medrxiv.org/content/10.1101/2020.05.18.20066902v1

8. Ly TDA, Zanini D, Laforge V, Arlotto S, Gentile S, Mendizabal H, Finaud M, Morel D, Quenette O, Malfuson-Clot-Faybesse P, Midejean A, Le-Dinh P, Daher G, Labarriere B, Morel-Roux AM, Coquet A, Augier P, Parola P, Chabriere E, Raoult D, Gautret P. Pattern of SARS-CoV-2 infection among dependant elderly residents living in long-term care facilities in Marseille, France, March-June 2020. Int J Antimicrob Agents. 2020 Dec;56(6):106219. doi: 10.1016/j.ijantimicag.2020.106219. Epub 2020 Nov 13. PMID: 33189890; PMCID: PMC7661959.

9. Alam MM, Mahmud S, Rahman MM, Simpson J, Aggarwal S, Ahmed Z. Clinical Outcomes of Early Treatment With Doxycycline for 89 High-Risk COVID-19 Patients in Long-Term Care Facilities in New York. Cureus. 2020;12(8): e9658. Published 2020 Aug 11. doi:10.7759/cureus.9658.

10. Cangiano B, Fatti LM, Danesi L, Gazzano G, Croci M, Vitale G, Gilardini L, Bonadonna S, Chiodini I, Caparello CF, Conti A, Persani L, Stramba-Badiale M, Bonomi M. Mortality in an Italian nursing home during COVID-19 pandemic: correlation with gender, age, ADL, vitamin D supplementation, and limitations of the diagnostic tests. Aging (Albany NY). 2020 Dec 22;12. doi: 10.18632/aging.202307. Epub ahead of print. PMID: 33353888.

11. Monica Leriger et al. (2020), American Senior Communities, Indianapolis, IN, personal communication, 2020. A Novel Study on the Use of Hydroxychloroquine in COVID-19 Positive Residents in a Nursing Home Setting

12. McCullough PA, Alexander, PE et al. Multifaceted highly targeted sequential multidrug treatment of early ambulatory high-risk SARS-CoV-2 infection (COVID-19). Reviews in Cardiovascular Medicine. 2020; 21 (4):517–530. DOI: 10.31083/j.rcm.2020.04.264.

13. Sodhi M, Etminan M. Therapeutic Potential for Tetracyclines in the Treatment of COVID-19. Pharmacotherapy. 2020 May;40(5):487–488. doi: 10.1002/phar.2395. Epub 2020 May 4. PMID: 32267566; PMCID: PMC7262278.

14. Roy SK, Kubiak BD, Albert SP, Vieau CJ, Gatto L, Golub L, Lee HM, Sookhu S, Vodovotz Y, Nieman GF. Chemically modified tetracycline 3 prevents acute respiratory distress syndrome in a porcine model of sepsis + ischemia/reperfusion-induced lung injury. Shock. 2012 Apr;37(4):424–32. doi: 10.1097/SHK.0b013e318245f2f9. PMID: 22258231.

15. Sadowsky D, Nieman G, Barclay D, Mi Q, Zamora R, Constantine G, Golub L, Lee HM, Roy S, Gatto LA, Vodovotz Y. Impact of chemically-modified tetracycline 3 on intertwined physiological, biochemical, and inflammatory networks in porcine sepsis/ARDS. Int J Burns Trauma. 2015 Mar 20;5(1):22–35. PMID: 26064799; PMCID: PMC4448085.

16. Golub LM, Elburki MS, Walker C, Ryan M, Sorsa T, Tenenbaum H, Goldberg M, Wolff M, Gu Y. Non-antibacterial tetracycline formulations: host-modulators in the treatment of periodontitis and relevant systemic diseases. Int Dent J. 2016 Jun;66(3):127–35. doi: 10.1111/idj.12221. Epub 2016 Mar 23. PMID: 27009489.

17. Gendrot M, Andreani J, Jardot P, Hutter S, Delandre O, Boxberger M, Mosnier J, Le Bideau M, Duflot I, Fonta I, Rolland C, Bogreau H, La Scola B, Pradines B. In Vitro Antiviral Activity of Doxycycline against SARS-CoV-2. Molecules. 2020 Oct 31;25(21):5064. doi: 10.3390/molecules25215064. PMID: 33142770; PMCID: PMC7663271.

18. Mosquera-Sulbaran JA, Hernández-Fonseca H. Tetracycline and viruses: a possible treatment for COVID-19? Arch Virol. 2020 Nov 2:1–7. doi: 10.1007/s00705-020-04860-8. Epub ahead of print. PMID: 33136210; PMCID: PMC7604546.

19. Yang W, Kandula S, Huynh M, Greene SK, Van Wye G, Li W, Chan HT, McGibbon E, Yeung A, Olson D, Fine A, Shaman J. Estimating the infection-fatality risk of SARS-CoV-2 in New York City during the spring 2020 pandemic wave: a model-based analysis. Lancet Infect Dis. 2020 Oct 19: S1473-3099(20)30769-6. doi: 10.1016/S1473-3099(20)30769-6. Epub ahead of print. Erratum in: Lancet Infect Dis. 2021 Jan;21(1): e1. PMID: 33091374; PMCID: PMC7572090.

20. Panagiotou OA, Kosar CM, White EM, Bantis LE, Yang X, Santostefano CM, Feifer RA, Blackman C, Rudolph JL, Gravenstein S, Mor V. Risk Factors Associated With All-Cause 30-Day Mortality in Nursing Home Residents With COVID-19. JAMA Intern Med. 2021 Jan 4. doi: 10.1001/jamainternmed.2020.7968. Epub ahead of print. PMID: 33394006.

21. Belmin J, Um-Din N, Donadio C, et al. Coronavirus Disease 2019 Outcomes in French Nursing Homes That Implemented Staff Confinement With Residents. JAMA Netw Open. 2020;3(8):e2017533. Published 2020 Aug 3. doi:10.1001/jamanetworkopen.2020.17533.

22. Davidson PM, Szanton SL. Nursing homes and COVID-19: We can and should do better. J Clin Nurs. 2020;29(15-16):2758–2759. doi:10.1111/jocn.15297.

23. Abrams HR, Loomer L, Gandhi A, Grabowski DC. Characteristics of U.S. Nursing homes with COVID-19 cases. J Am Geriatr Soc. 2020; 68:1653–56. https://doi.org/10.1111/jgs.16661 PMID:32484912.

24. Graham NSN, Junghans C, Downes R, Sendall C, Lai H, McKirdy A, Elliott P, Howard R, Wingfield D, Priestman M, Ciechonska M, Cameron L, Storch M, Crone MA, Freemont PS, Randell P, McLaren R, Lang N, Ladhani S, Sanderson F, Sharp DJ. SARS-CoV-2 infection, clinical features and outcome of COVID-19 in United Kingdom nursing homes. J Infect. 2020 Sep;81(3):411–419. doi: 10.1016/j.jinf.2020.05.073. Epub 2020 Jun 3. PMID: 32504743.

25. McMichael TM, Currie DW, Clark S, Pogosjans S, Kay M, Schwartz NG, Lewis J, Baer A, Kawakami V, Lukoff MD, Ferro J, Brostrom-Smith C, Rea TD, Sayre MR, Riedo FX, Russell D, Hiatt B, Montgomery P, Rao AK, Chow EJ, Tobolowsky F, Hughes MJ, Bardossy AC, Oakley LP, Jacobs JR, Stone ND, Reddy SC, Jernigan JA, Honein MA, Clark TA, Duchin JS; Public Health–Seattle and King County, Evergreen Health, and CDC COVID-19 Investigation Team. Epidemiology of Covid-19 in a Long-Term Care Facility in King County, Washington. N Engl J Med. 2020 May 21;382(21):2005–2011. doi: 10.1056/NEJMoa2005412. Epub 2020 Mar 27. PMID: 32220208; PMCID: PMC7121761.

26. Tan LF, Seetharaman SK. COVID-19 Outbreak in Nursing Homes in Singapore. J Microbiol Immunol Infect. 2020 May 13. doi: 10.1016/j.jmii.2020.04.018. Epub ahead of print. PMID: 32405290; PMCID: PMC7219412.

27. Arons MM, Hatfield KM, Reddy SC, Kimball A, James A, Jacobs JR, Taylor J, Spicer K, Bardossy AC, Oakley LP, Tanwar S, Dyal JW, Harney J, Chisty Z, Bell JM, Methner M, Paul P, Carlson CM, McLaughlin HP, Thornburg N, Tong S, Tamin A, Tao Y, Uehara A, Harcourt J, Clark S, Brostrom-Smith C, Page LC, Kay M, Lewis J, Montgomery P, Stone ND, Clark TA, Honein MA, Duchin JS, Jernigan JA; Public Health–Seattle and King County and CDC COVID-19 Investigation Team. Presymptomatic SARS-CoV-2 Infections and Transmission in a Skilled Nursing Facility. N Engl J Med. 2020 May 28;382(22):2081–2090. doi: 10.1056/NEJMoa2008457. Epub 2020 Apr 24. PMID: 32329971; PMCID: PMC7200056.

28. Kemenesi G, Kornya L, Tóth GE, Kurucz K, Zeghbib S, Somogyi BA, Zöldi V, Urbán P, Herczeg R, Jakab F. Nursing homes and the elderly regarding the COVID-19 pandemic: situation report from Hungary. Geroscience. 2020 May 18;42(4):1–7. doi: 10.1007/s11357-020-00195-z. Epub ahead of print. PMID: 32426693; PMCID: PMC7232926.

29. Burki T. England and Wales see 20L000 excess deaths in care homes. Lancet. 2020 May 23;395(10237):1602. doi: 10.1016/S0140-6736(20)31199-5. PMID: 32446403; PMCID: PMC7241982.

30. Tse MM, Pun SP, Benzie IF. Experiencing SARS: perspectives of the elderly residents and health care professionals in a Hong Kong nursing home. Geriatr Nurs. 2003; 24:266–69. https://doi.org/10.1016/s0197-4572(03)00251-9 PMID:14571239.

31. COVID-19 data in Quebec. url: https://www.inspq.qc.ca/covid-19/donnees (Accessed on December 24th 2020).

32. COVEXIT. url: https://covexit.com/covid-19-in-nursing-homes-a-way-forward-to-end-the-tragedy/ (Accessed on December 25th 2020).

33. Public Health Ontario. COVID-19 in Ontario: January 15, 2020 to January 10, 2021. url: https://www.publichealthontario.ca/-/media/documents/ncov/epi/2020/covid-19-daily-epi-summary-report.pdf?la=en (Accessed on January 11th 2021).

34. The Chief Public Health Officer of Canada’s Report on the State of Public Health in Canada 2020. COVID-19 in Canada. url: https://www.canada.ca/content/dam/phac-aspc/documents/corporate/publications/chief-public-health-officer-reports-state-public-health-canada/from-risk-resilience-equity-approach-covid-19/cpho-covid-report-eng.pdf (Accessed on January 11th 2021).

35. CDC. Characterization of COVID-19 in Assisted Living Facilities — 39 States, October 2020. MMWR. url: https://www.cdc.gov/mmwr/volumes/69/wr/mm6946a3.htm (Accessed December 25th 2020).

36. Almost half of US COVID-19 deaths are linked to nursing homes. url: https://nypost.com/2020/06/27/almost-half-of-us-covid-19-deaths-are-linked-to-nursing-homes/ (Accessed on December 24th 2020).

37. Coronavirus deaths in NY nursing homes higher than state data shows, feds say. url: https://nypost.com/2020/08/11/covid-deaths-in-ny-nursing-homes-higher-than-data-shows-feds/ (Accessed on December 25th 2020).

38. McCullough PA. Favipiravir and the Need for Early Ambulatory Treatment of SARS-CoV-2 Infection (COVID-19). Antimicrob Agents Chemother. 2020 Nov 17;64(12):e02017–20. doi: 10.1128/AAC.02017-20. PMID: 32967849; PMCID: PMC7674042.

39. Rajter JC, Sherman MS, Fatteh N, Vogel F, Sacks J, Rajter JJ. Use of Ivermectin Is Associated With Lower Mortality in Hospitalized Patients With Coronavirus Disease 2019: The Ivermectin in COVID Nineteen Study. Chest. 2021 Jan;159(1):85–92. doi: 10.1016/j.chest.2020.10.009. Epub 2020 Oct 13. PMID: 33065103; PMCID: PMC7550891.

40. Front Line COVID-19 Critical Care Alliance. FLCCC. url: https://covid19criticalcare.com/ (Accessed on January 15th 2021).

41. Ansarin K, Tolouian R, Ardalan M, Taghizadieh A, Varshochi M, Teimouri S, Vaezi T, Valizadeh H, Saleh P, Safiri S, Chapman KR. Effect of bromhexine on clinical outcomes and mortality in COVID-19 patients: A randomized clinical trial. Bioimpacts. 2020;10(4):209–215. doi: 10.34172/bi.2020.27. Epub 2020 Jul 19. PMID: 32983936; PMCID: PMC7502909.

42. Maggio R, Corsini GU. Repurposing the mucolytic cough suppressant and TMPRSS2 protease inhibitor bromhexine for the prevention and management of SARS-CoV-2 infection. Pharmacol Res. 2020 Jul;157:104837. doi: 10.1016/j.phrs.2020.104837. Epub 2020 Apr 22. PMID: 32334052; PMCID: PMC7175911.

43. Frieden TR. Evidence for Health Decision Making - Beyond Randomized, Controlled Trials. N Engl J Med. 2017 Aug 3;377(5):465–475. doi: 10.1056/NEJMra1614394. PMID: 28767357.

44. Public Law. ‘21st Century Cures Act. url: file:///C:/Users/Paul/Downloads/21%20Century%20Cures%20Act%20PLAW-114publ255%20(1).pdf (Accessed on January 25th 2021).

45. Cadegiani et al., medRxiv, doi:10.1101/2020.10.31.20223883 (Preprint). Early COVID-19 Therapy with Azithromycin Plus Nitazoxanide, Ivermectin or Hydroxychloroquine in Outpatient Settings Significantly Reduced Symptoms Compared to Known Outcomes in Untreated Patients.

46. Early outpatient treatment: ‘Essential part of COVID-19 solution’. Senator Ron Johnson, US Senate. url: https://www.israelnationalnews.com/News/News.aspx/291718 (Accessed on January 23rd 2021).

47. Senate Hearing on COVID-19 Outpatient Treatment. url: https://www.c-span.org/video/?478159-1/senate-hearing-covid-19-outpatient-treatment (Accessed on January 25th 2021).

